# Plasma and CSF neurofilament light chain measured by Simoa and Lumipulse: an inter-platform comparison across neurological disorders

**DOI:** 10.64898/2026.06.01.26354573

**Authors:** Andrea Toja, Virginia Quaresima, Chiara Tolassi, Tommaso Merati, Chiara Trasciatti, Simona G. Signorini, Andrea Morotti, Francesco Berinato, Loris Poli, Liberato Stabile, Irene Girotto, Marianna Bertoni, Cinzia Zatti, Alessandro Magliozzi, Caterina Martinuzzo, Claudia Pangrazio, Klaudia Eshja, Giulia Foresti, Ilenia Libri, Eqrem Rusi, Marta Bianchi, Viviana Cristillo, Irene Volonghi, Alice Galli, Andrea Rizzardi, Salvatore Caratozzolo, Chiara Agosti, Rosanna Colao, Carmelo Rodolico, Elena Marcello, Fabrizio Gardoni, Monica Di Luca, Henrik Zetterberg, Nicholas J. Ashton, Duilio Brugnoni, Andrea Pilotto, Alessandro Padovani

## Abstract

**Introduction:** Blood neurofilament light chain (NfL) is an accessible biomarker of neuroaxonal injury across a broad range of neurological disorders, but its clinical implementation requires robust cross-platform analytical and clinical comparability. The objective of this study was to evaluate the analytical and clinical comparability of plasma NfL measurements using Simoa and Lumipulse across different neurological conditions, by assessing cross-platform agreement and the ability of both assays to distinguish neurological diseases from healthy controls. Paired CSF analyses were performed in a subset of participants to biologically anchor plasma findings to the central compartment.

**Methods:** 383 individuals were included, comprising healthy controls and patients with neurodegenerative conditions, multiple sclerosis and stroke. Plasma NfL was measured in all participants using both Simoa and Lumipulse, with paired CSF analyses in a subset of 92 individuals. The Lumipulse testing intermediate precision and between-day repeatability was assessed as by the CLSI EP15. Cross-platform agreement for plasma NfL was evaluated using correlation analyses, Passing–Bablok regression and Bland–Altman analysis. Associations between plasma/CSF NfL concentrations were assessed using Spearman’s rank correlation analysis for each platform, separately. Age-adjusted cross-diagnostic differences were evaluated using permutation ANCOVA and multiple linear regression models for each platform, separately.

**Results:** Plasma NfL measured by Simoa and Lumipulse showed strong cross-platform concordance in the whole cohort (ρ=0.90), with similarly strong concordance observed for CSF NfL in the subset with paired samples (ρ=0.90). Method-comparison analyses in plasma demonstrated consistent agreement between platforms, with identifiable constant and proportional bias, alongside systematically higher absolute plasma NfL values measured by Lumipulse. Within-platform analyses showed significant correlations between plasma and CSF NfL concentrations (ρ=0.72 for Simoa; ρ=0.78 for Lumipulse). Noteworthy, Lumipulse NfL CSF and Blood kits exhibited high precision and analytical accuracy. Across both assays, plasma NfL increased with age and was significantly elevated in patients with neurological disorders compared with healthy controls.

**Discussion:** Simoa and Lumipulse capture a consistent biological signal in plasma across patients with neurological disorders, although their absolute NfL values differ, supporting the use of platform-specific reference ranges in clinical practice.

## Introduction

Blood neurofilament light chain (NfL) is widely recognized as a biomarker of non-specific axonal injury across several neurological disorders [1,2]. NfL is a structural component of neuronal cytoskeletal neurofilaments, which are localized in the axonal cytoplasm and contribute to axonal stability and radial growth [1,3]. When axonal damage occurs, e.g. due to neurodegenerative, inflammatory, traumatic, or vascular mechanisms within the central nervous system (CNS), NfL is released into the extracellular space and subsequently detectable in both cerebrospinal fluid (CSF) and blood [1,3–10]. Because blood concentrations are substantially lower than those in CSF [11], the development of ultrasensitive analytical methods has been crucial for its reliable quantification. The possibility of measuring NfL in blood has provided a valuable opportunity to quantify axonal damage using an easily accessible biological matrix, thereby also allowing for longitudinal assessments. Accordingly, blood NfL has emerged as a promising axonal injury biomarker across heterogeneous neurological conditions. However, its interpretation is not straightforward, as concentrations are also strongly influenced by age [12,13], which must be taken into account when comparing individuals or diagnostic groups.

Consistency across measurement platforms is highly relevant to the broader clinical implementation of blood NfL. Among these methods, single-molecule array (Simoa) has become one of the most widely used platforms for blood NfL quantification and has shown high concordance with CSF NfL levels [11]. However, its use in routine clinical laboratory settings remains limited. Chemiluminescent enzyme immunoassays (CLEIA) designed for fully automated platforms have been developed for the measurement of NfL in both CSF and blood, showing good correlation with Simoa-based assays [14]. Correlation alone does not ensure clinical comparability between platforms, highlighting the need for robust cross-platform validation across different neurological disorders to support clinical application. Therefore, plasma and CSF NfL measured by Simoa and Lumipulse were directly compared in a large, clinically heterogeneous cohort of healthy controls and patients with neurodegenerative, inflammatory, and vascular neurological disorders, including Alzheimer’s disease (AD), dementia with Lewy bodies (DLB), frontotemporal dementia (FTD), motor neuron disease (MND), multiple sclerosis (MS), and ischemic stroke. The study aimed to assess analytical and clinical cross-platform comparability by evaluating agreement between platforms and the ability of both assays to distinguish neurological diseases from healthy controls. In addition, Lumipulse testing robustness and analytical performance was evaluated on both plasma and CSF reagent kits.

## Materials and methods

### Study population

The samples included in this study were retrospectively selected from a cohort of 383 individuals who underwent plasma collection at the department of S.C Neurology of ASST Spedali Civili Hospital, Italy, as part of an ongoing prospective study on plasma biomarkers. Patients were diagnosed as AD, DLB, FTD, MND, MS and ischemic stroke according to the current diagnostic criteria [15–19]. AD diagnosis was carried out in 139 patients clinically and confirmed biologically according to CSF hybrid ratio Aβ_42_/p-tau181 ratio <11.1 [20–22]. A group of n=88 neurologically and cognitively normal individuals (HC) was recruited from participants’ caregivers, as part of the Life-BIO cohort. The following exclusion criteria were applied: (i) diagnosis of any neurological disorder; (ii) presence of subjective cognitive complaints; (iii) abnormal neurological examination and Montreal Cognitive Assessment screening; (iv) major psychiatric disorder; or (v) recent inflammatory events. The study was approved by the local ethics committee (NP 1471, DMA, Brescia) and Neuromultibio Study (NP5285, approved by the Local Brescia ASST Ethic Committee on the 10.05.2022) and performed in conformity with the Declaration of Helsinki; informed consent was obtained from each study participant or their legally authorized representative.

### Plasma collection and analysis

Venous blood was collected into 4.9 ml tubes containing K2-ethylenediaminetetraacetic acid (K2-EDTA, S-Monovette tubes Sarstedt, #04.1932.001). Following 5–10 gentle inversions, samples were centrifuged at 2500 g for 10 min at room temperature [23]. Plasma was subsequently aliquoted into 0.5 mL polypropylene cryotube aliquots (ClearLine, #CL2ARBIPSTS) and stored at −80°C until Lumipulse and SIMOA analysis.

### CSF collection and analysis

A subset of patients underwent lumbar puncture in fasting condition according to the standardized protocol of the outpatient neurodegenerative clinic. CSF specimens (15 mL) were collected into sterile polypropylene tubes (CSF tubes/S-Monovette®, #63.504.027) and gently mixed to avoid gradient effects, following international guidelines for Alzheimer’s disease (AD) biomarker handling [24,25]. Routine assessments and core AD biomarker tests were performed using Lumipulse G600 II platform, applying the internal Aβ_42_/p-tau181 ratio cut-off of <11.1 [20].

Residual CSF volumes were aliquoted into 0.7 mL low binding polypropylene tubes (Fluidx, #783931) for biorepository purposes or into 1.5 ml tubes (CLEARLine, #CL754/S) for potential re-testing. All aliquots were subsequently frozen at ultra-low temperature freezing (ULTF) -80°C for long-term-storage.

A subset of N=92 patients (44 AD, 9 DLB, 10 FTD, 14 MND, 15 MS) underwent CSF NfL testing using both Lumipulse and Simoa, with the sampling interval between plasma and CSF ranging from −61 to +61 days.

### SIMOA and Lumipulse Analysis

Sample preparation and analysis adhere to the 2025 NACC ADRC Biofluid Biomarker Best Practices and the Alzheimer’s Association international guidelines [24,25]. To ensure protein integrity, particularly given that NfL remains stable in neat CSF for less than 24 hours at room temperature, all CSF aliquots were immediately frozen at -80°C upon collection.

On the day of analysis, plasma and CSF specimens were thawed and equilibrated to room temperature (21–23 °C). Following the manufacturer’s protocol, plasma samples were centrifuged at 2000g for 5 minutes, while CSF samples were vortexed and processed without further centrifugation, except in cases of visible blood contamination.

SIMOA analyses were performed on SR-X platform with commercially available Neuro 2-Plex B Advantage Kit Simoa® (Quanterix, Billerica MA) at the Neurobiorepository and Laboratory of advanced biological markers, University of Brescia and ASST Spedali Civili Hospital (Brescia, Italy). The Neurology 2-Plex B assay targets simultaneously neurofilament light (NfL) and glial fibrillary acidic protein (GFAP) in both CSF and blood samples. Each sample, for each analyte, was analysed in technical duplicate with a CV< 20%. In plasma the functional Lower Limit of Quantification (fLLOQ) is 1.60 pg/mL for NfL; the Limit of Detection (LOD) is 0.012-0.149 pg/mL for NfL; the assay dynamic range is 0-2000 pg/mL for NfL. In CSF the fLLOQ and the Upper Limit of Quantification (ULOQ) is 10X than plasma.

To minimize pre-analytical variability and prevent material loss, the majority of both CSF and plasma samples for Lumipulse testing were analyzed directly within their primary storage tubes (ClearLine, #CL2ARBIPSTS). However, for samples with insufficient volume, plasma and CSF specimens were transferred into Hitachi sample cups to ensure compatibility with the instrument’s dead volume requirements.

All measurements were performed on the automated Lumipulse G600II platform (Fujirebio) using the following immunoreaction cartridges: Lumipulse G NfL Blood (REF: 81215; LOT# 5078, 5109) for plasma (Calibrators REF: 81422, Controls REF: 81421), and Lumipulse G NfL CSF (REF: 81426; LOT# 5078; Calibrators REF: 81413, Controls REF: 81414) for CSF samples.

### Precision assessment

The within-laboratory precision of the Lumipulse G600II NfL assays was evaluated for both plasma and CSF kits through a 5×5 repeated inter-day testing protocol, in accordance with the CLSI EP15-A3 guidelines and established validation frameworks [26,27]. To assess intermediate precision and between-run repeatability, ready-to-use quality control (QC) materials were used to ensure standardized testing performance.

For the plasma NfL assay, two levels of commercial QC samples (low - Plasma Level 1, PL1 and high – Plasma Level 2, PL2) were analyzed in five replicates per day over five consecutive days (25 independent runs). For the CSF NfL assay, the same 5×5 scheme was applied using three levels of commercial QC materials (CSF Level 1, CL1; CSF Level 2, CL2; CSF Level 3, CL3). During the assessment period, all commercial QC materials were stored at 4°C as per the manufacturer’s instructions. Intermediate precision and between-run coefficients of variation (%CV) were subsequently calculated to validate the analytical robustness of the Lumipulse G platform for both cartridges’ kits.

### Workflow total imprecision

To assess the assay’s robustness in a real-life clinical setting, we evaluated the impact of pre-analytical handling and storage conditions. A total of 67 CSF samples were analyzed in duplicate across two independent experimental runs. Samples underwent two distinct sampling and storage pathways: the first set of aliquots was managed and retrieved from the the Neurobiorepository (University of Brescia), while the second set was processed through the routine workflow of the Clinical Chemistry Laboratory (ASST Spedali Civili General Hospital of Brescia). Although the same analytical kit and platform were used for all runs, testing was performed at different time points and using different storage vials to assess internal method robustness and the influence of sample handling protocols. This approach provides a measure of assay reliability that goes beyond standard manufacturer-provided pre-commercial validation data, reflecting the expected variability in routine clinical practice.

### Statistical analyses

#### Distribution assessment

Normality of plasma and CSF NfL concentrations measured by the Simoa and Lumipulse platforms was assessed using the Shapiro–Wilk test on both raw (pg/mL) and natural log-transformed values. Distributional assumptions were further evaluated visually using quantile–quantile plots and histograms for each analytical platform.

#### Workflow total imprecision

The cumulative precision was estimated using the Root Mean Square (RMS) method, as recommended by Jones & Payne [28,29], to provide a single representative coefficient of variation (CV%) that accounts for variance across the assay’s dynamic range. In order to obtain a comprehensive estimate of the platform’s analytical performance, the Total Error (TE) was calculated by combining this cumulative precision profile with the empirical systematic percentage bias obtained from the Bland–Altman agreement analysis, with a 1.65 Z-score for a 95% confidence limit as by the following formula: TE = |Bias%| + 1.65 x CV% [30].

To further assess the agreement between replicates, Passing–Bablok regression was performed to detect potential constant or proportional biases, while a Bland–Altman analysis was used to visualize the distribution of differences and confirm the absence of concentration-dependent systematic errors. This integrated approach ensures that the measurement imprecision remains stable and clinically negligible across the entire spectrum of CSF NfL concentrations.

#### Cross-platform and CSF analyses

The association between plasma NfL concentrations measured by Lumipulse and Simoa was assessed using Spearman’s rank correlation coefficient in the overall cohort and within each diagnostic category. Method agreement was evaluated using Passing–Bablok regression and Bland–Altman analysis. Differences between paired plasma NfL measurements obtained with the two assay platforms were assessed using the Wilcoxon signed-rank test and t-test. In the subset of participants with paired CSF measurements, the association between plasma and CSF NfL concentrations was evaluated separately for the Lumipulse and Simoa platforms using Spearman’s rank correlation coefficient. Concordance between CSF NfL concentrations measured with Lumipulse and Simoa was also assessed using Spearman’s rank correlation coefficient.

#### Age-adjusted clinical analyses

To assess clinically relevant differences in plasma NfL across diagnostic categories while accounting for age, separate permutation-based ANCOVA models were fitted for Lumipulse and Simoa, using log-transformed plasma NfL as the dependent variable and diagnosis, age, and their interaction as predictors. As a sensitivity analysis, separate multiple linear regression models were fitted for Lumipulse and Simoa to examine the independent association of age and diagnostic groups with plasma NfL concentrations. In these models, natural log-transformed plasma NfL concentration was entered as the dependent variable, whereas age and diagnosis were included as independent variables. Diagnosis was treated as a categorical predictor, with HC as the reference category.

A two-sided significance level of 0.05 was adopted for all statistical tests. All analyses were conducted using R statistical software version 4.5.1.

## Results

### Study population and distribution of plasma and CSF NfL

N=139 patients were diagnosed as AD, N=26 as DLB, N=24 as FTD, n=37 as MND, N=30 as MS, N=39 as ischemic stroke. Demographic and clinical characteristics of the study population are summarized in Table 1. Plasma neurofilament light chain (NfL) concentrations measured by both Simoa and Lumipulse showed a markedly non-normal distribution on raw values (Shapiro–Wilk: Simoa W = 0.58, p < 0.001; Lumipulse W = 0.52, p < 0.001). After log-transformation, the distribution of Simoa values improved symmetry, although normality was not fully achieved (W = 0.99, p = 0.01), whereas Lumipulse values remained significantly non-normal (W = 0.92, p < 0.001). Visual inspection using Q–Q plots and histograms confirmed a right-skewed distribution on raw values and an overall improvement after log-transformation (Supplementary Figure 1).

**Table 1.** Clinical and demographic characteristics of the cohort.

|  | *N* | *Sex (M/F)* | *Age (years)* | **NfL Simoa (pg/mL)** |  | **NfL Lumipulse (pg/mL)** |  |
| --- | --- | --- | --- | --- | --- | --- | --- |
|  |  |  |  | *Mean ± SD* | *Median (IQR)* | *Mean ± SD* | *Median (IQR)* |
| **HC** | 88 | 30/58 | 58.7 ± 14.6 | 8.6 ± 4.6 | 8.0 (5.3–10.6) | 19.2 ± 8.8 | 18.3 (13.7–22.4) |
| **AD** | 139 | 54/85 | 71.4 ± 7.8 | 28.6 ± 33.6 | 21.8 (16.6–30.5) | 36.8 ± 40.5 | 28.7 (23.5–35.9) |
| **DLB** | 26 | 18/8 | 76.1 ± 5.7 | 29.8 ± 20.5 | 23.4 (17.0–38.8) | 42.6 ± 38.1 | 31.5 (22.9–45.2) |
| **FTD** | 24 | 15/9 | 69.4 ± 6.7 | 30.8 ± 24.2 | 24.0 (15.5–35.5) | 42.5 ± 31.8 | 33.8 (22.7–51.1) |
| **MND** | 37 | 20/17 | 68.8 ± 9.0 | 79.5 ± 56.0 | 58.4 (38.4–121.2) | 110.3 ± 77.4 | 88.8 (55.4–153.3) |
| **STROKE** | 39 | 18/21 | 72.6 ± 7.2 | 47.9 ± 60.3 | 31.7 (19.1–52.3) | 69.3 ± 92.1 | 44.0 (25.7–69.2) |
| **MS** | 30 | 6/24 | 31.1 ± 12.4 | 19.5 ± 29.3 | 10.6 (5.7–15.1) | 39.7 ± 60.1 | 20.8 (14.8–29.3) |

HC, healthy controls; AD, Alzheimer's disease; DLB, dementia with Lewy bodies, FTD, frontotemporal dementia; MND, motor neuron disease; STROKE, ischemic stroke; MS, multiple sclerosis.

In the subset of individuals with available CSF measurements (n = 92), CSF NfL concentrations measured using both Lumipulse and Simoa also showed a markedly right-skewed distribution on raw values (Shapiro–Wilk: Lumipulse W = 0.72, p < 0.001; Simoa W = 0.74, p < 0.001). Log-transformation improved the symmetry of the distributions (Lumipulse W = 0.97, p = 0.046; Simoa W = 0.97, p = 0.057), with Q–Q plots confirming a substantial reduction in skewness after transformation (Supplementary Figure 2).

### Precision and repeatability

High stability was shown by NfL plasma levels, with a CV of 3.99% and 3.26% for PL1 and PL2 respectively (Supplementary Tables 1, 2 and 3). Likewise, the %CVs for the three aliquots to verify CSF kit performance were 3.35, 2.68 and 2.61, for the low (CL1), medium (CL2) and high (CL3) levels respectively (Supplementary Tables 3, 4, 5 and 6).

### Workflow total imprecision

To evaluate the analytical robustness of the assay in a clinical real-life, an extensive precision study was conducted on 67 CSF samples across a wide dynamic range. The inter-assay precision, estimated using the RMS method [29], yielded a CV of 7.46% (95% CI: 6.39% – 8.98%).

In order to obtain a realistic estimate of the platform’s performance, a Bland–Altman analysis was first applied to calculate the empirical systematic percentage bias between replicates, which was established at -4.33% (95% CI: -6.70% to -1.97%; Supplementary Table 7, Supplementary Figure 3); the calculated TE was 16.64% [30].

This analytical performance is consistent with the current international consensus for neurodegenerative CSF biomarkers, which report an intra-laboratory coefficient of variation ranging between 5% and 19% [31]. Achieving a CV close to the lower limit of this reference range suggests the high reproducibility for the platform when operating within routine conditions.

The assay’s overall stability was further validated through Passing–Bablok regression, which demonstrated the absence of proportional bias (slope: 1.02; 95% CI: 0.99 – 1.06) and constant systematic deviation (intercept: 19.58 pg/mL; 95% CI: -10.57 – 50.00; Supplementary Table 8, Supplementary Figure 4).

### Concordance between Lumipulse and Simoa plasma NfL

Plasma neurofilament light chain (NfL) concentrations measured by Lumipulse and Simoa were strongly correlated in the overall cohort on both raw and log-transformed values (Spearman’s ρ = 0.90, p < 0.001 both; Figure 1). When stratified by diagnostic category, strong correlations between Lumipulse and Simoa plasma NfL were observed across all groups (Figure 2 and Supplementary Figure 5). Spearman’s correlation coefficients ranged from 0.70 in healthy controls to 0.97 in patients with stroke, with all associations reaching statistical significance (all p < 0.001).

**Figure 1.**
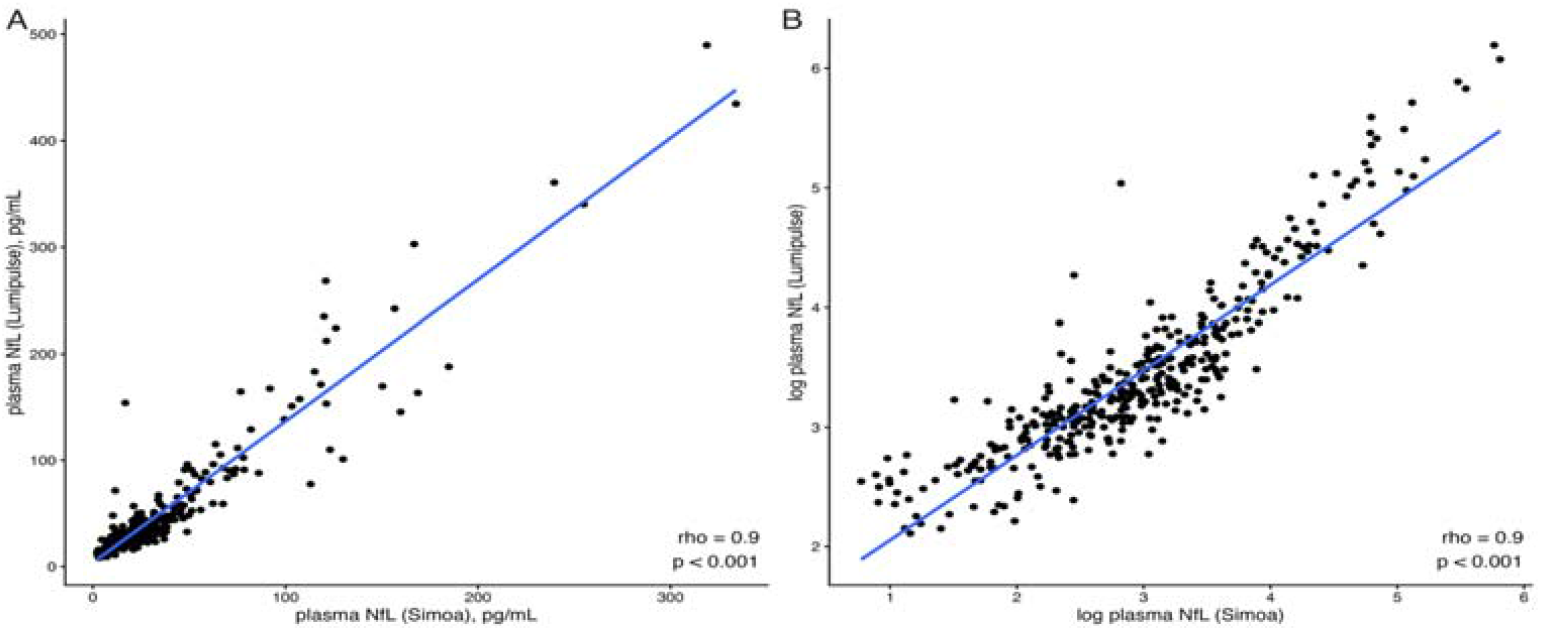
Correlation between plasma neurofilament light chain (NfL) measured by Lumipulse and Simoa. (A) Scatterplot of raw plasma NfL concentrations (pg/mL) measured by Lumipulse versus Simoa. (B) Scatterplot of natural log-transformed plasma NfL concentrations. In both panels, the solid line represents the linear regression fit for visualization purposes. Spearman’s rank correlation coefficient (ρ) and p value are reported within each panel. Log-transformed values are shown to improve data visualization.

**Figure 2.**
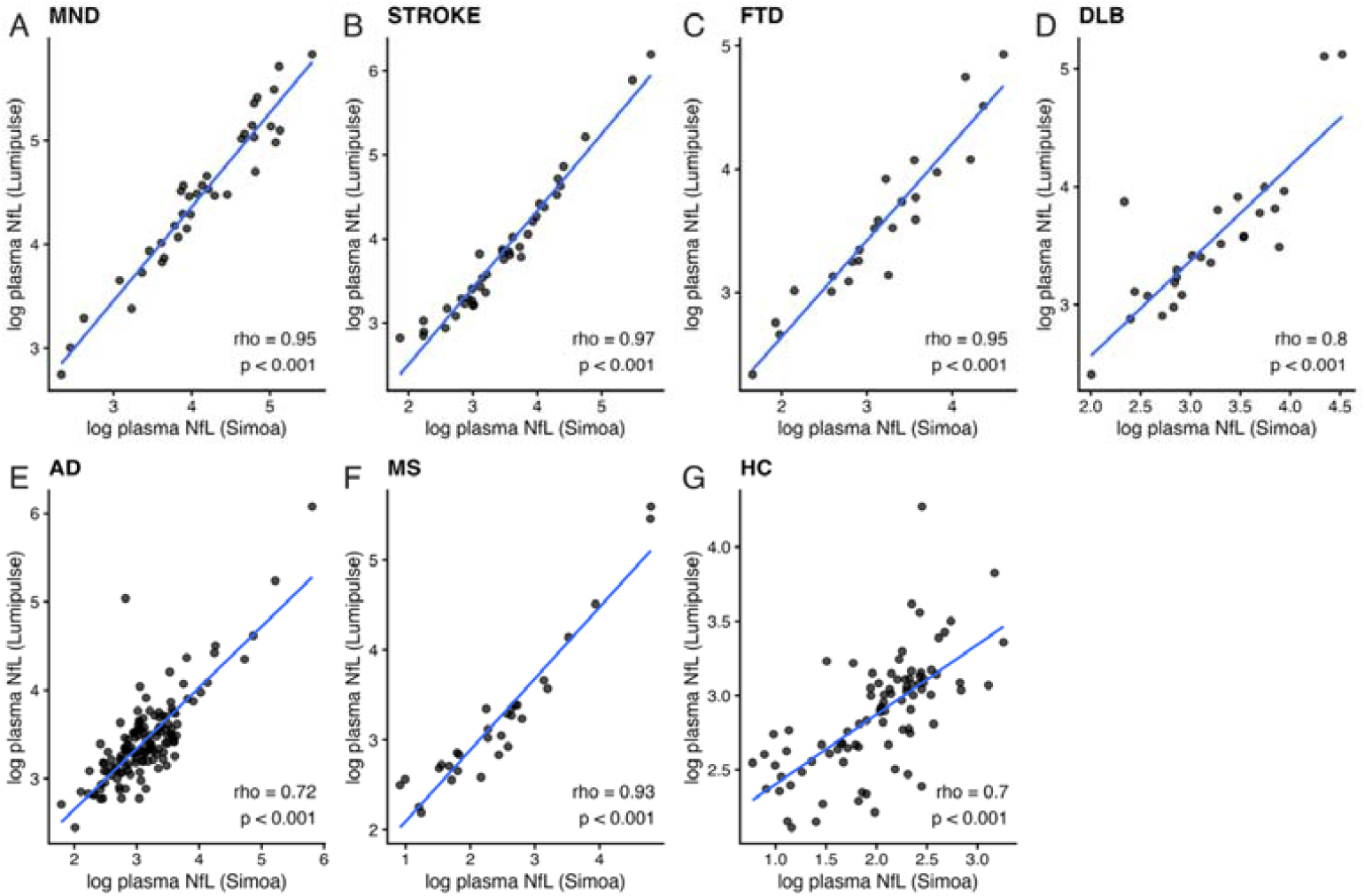
Spearman correlation between plasma neurofilament light chain (NfL) measured by Lumipulse and Simoa within diagnostic categories (log-transformed values) Multipanel scatterplots showing the association between natural log-transformed plasma NfL concentrations measured by Lumipulse and Simoa in (A) motor neuron disease (MND), (B) ischemic stroke (STROKE), (C) frontotemporal dementia (FTD), (D) dementia with Lewy bodies (DLB), (E) Alzheimer’s disease (AD), (F) multiple sclerosis (MS), and (G) healthy controls (HC). Spearman’s rank correlation coefficient (ρ) and p value are reported within each panel; the solid line indicates the least-squares fit for visualization.

### Method comparison: Passing-Bablok and Bland-Altman analyses

Passing–Bablok regression analysis showed a slope of 1.18 (95% CI 1.12–1.25) and an intercept of 6.82 pg/mL (95% CI 5.42–7.82), indicating a proportional and constant positive bias of Lumipulse relative to Simoa (Figure 3). Bland–Altman analysis showed a mean bias of 13.8 pg/mL, with 95% limits of agreement ranging from −29.2 to 56.8 pg/mL (Figure 4). A systematic difference between methods was confirmed by both the one-sample t-test and the Wilcoxon signed-rank test (both p < 0.001).

**Figure 3.**
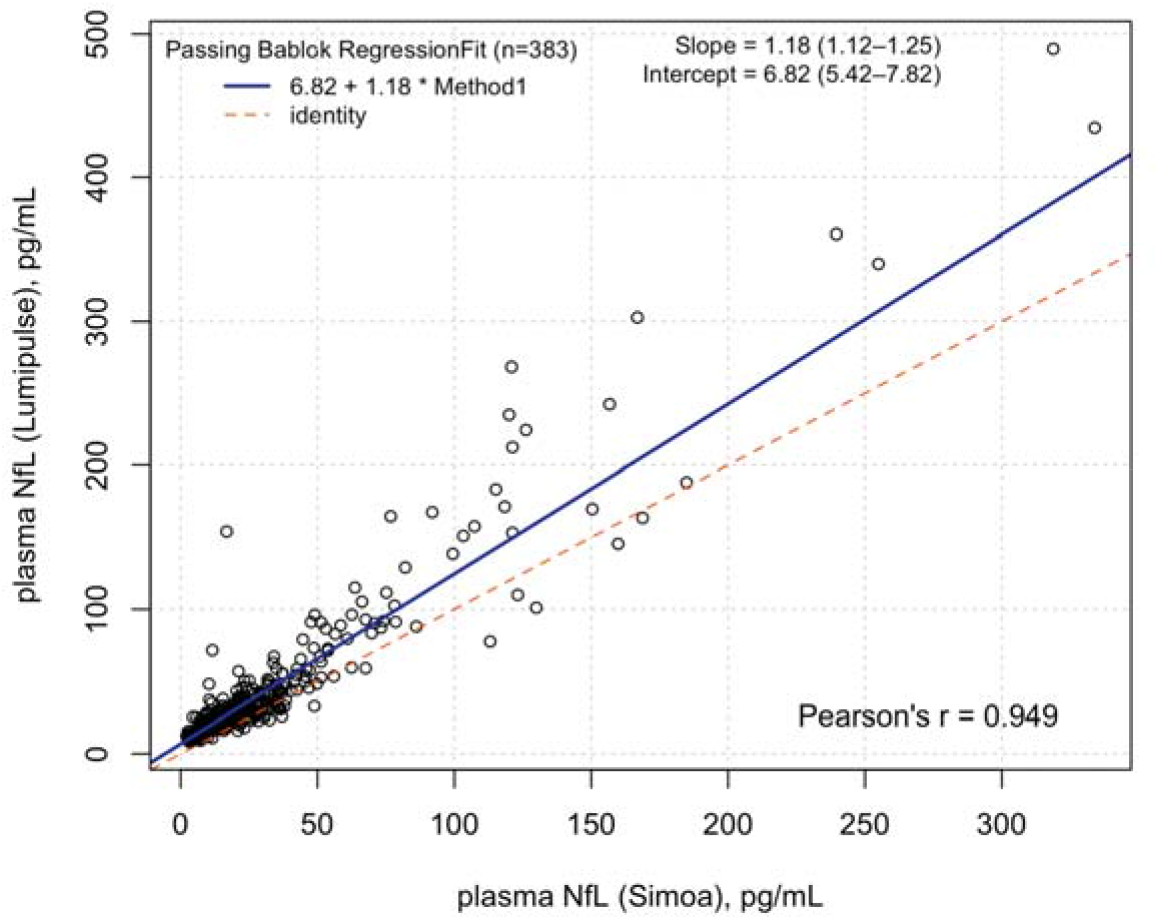
Passing–Bablok regression comparing plasma neurofilament light chain (NfL) measured by Lumipulse and Simoa. Passing–Bablok regression of plasma NfL concentrations. The solid line represents the regression fit, and the dashed line indicates the line of identity (y = x). Values in brackets represent confidence interval.

**Figure 4.**
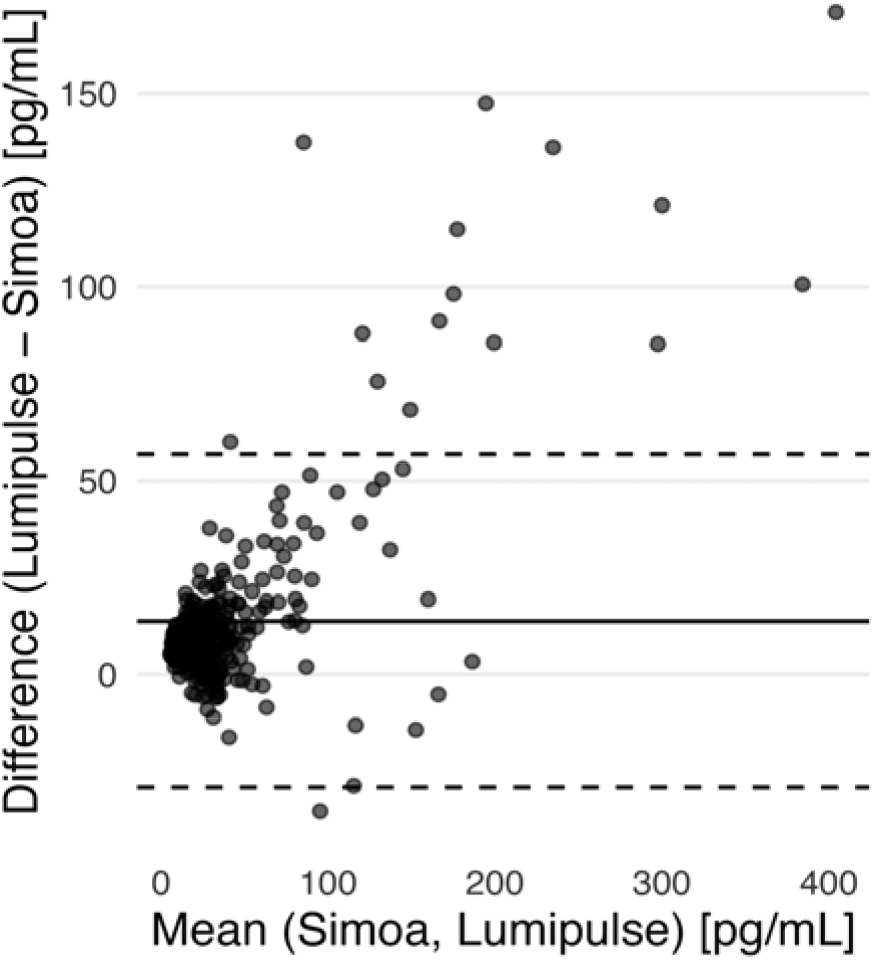
Bland–Altman plot comparing plasma neurofilament light chain (NfL) measured by Lumipulse and Simoa. Differences between methods (Lumipulse − Simoa, pg/mL) are plotted against the mean of the two measurements. The solid horizontal line represents the mean bias, and the dashed lines indicate the 95% limits of agreement (bias ±1.96 SD). The solid diagonal line represents the linear regression used to assess proportional bias.

### CSF-plasma NfL concordance using SIMOA and LUMIPULSE analyses

In the subset with paired plasma and CSF measurements collected within a −61 to +61 day interval (n = 92), significant correlations were observed between plasma and CSF NfL concentrations for both analytical platforms. Plasma NfL measured with Lumipulse showed a strong correlation with Lumipulse CSF NfL concentrations (Spearman’s ρ = 0.78, p < 0.001). Similarly, plasma NfL measured with Simoa correlated significantly with Simoa CSF NfL concentrations (Spearman’s ρ = 0.72, p < 0.001). A very strong correlation was observed between CSF NfL concentrations measured with Lumipulse and Simoa (Spearman’s ρ = 0.90, p < 0.001;Figure 5).

**Figure 5.**
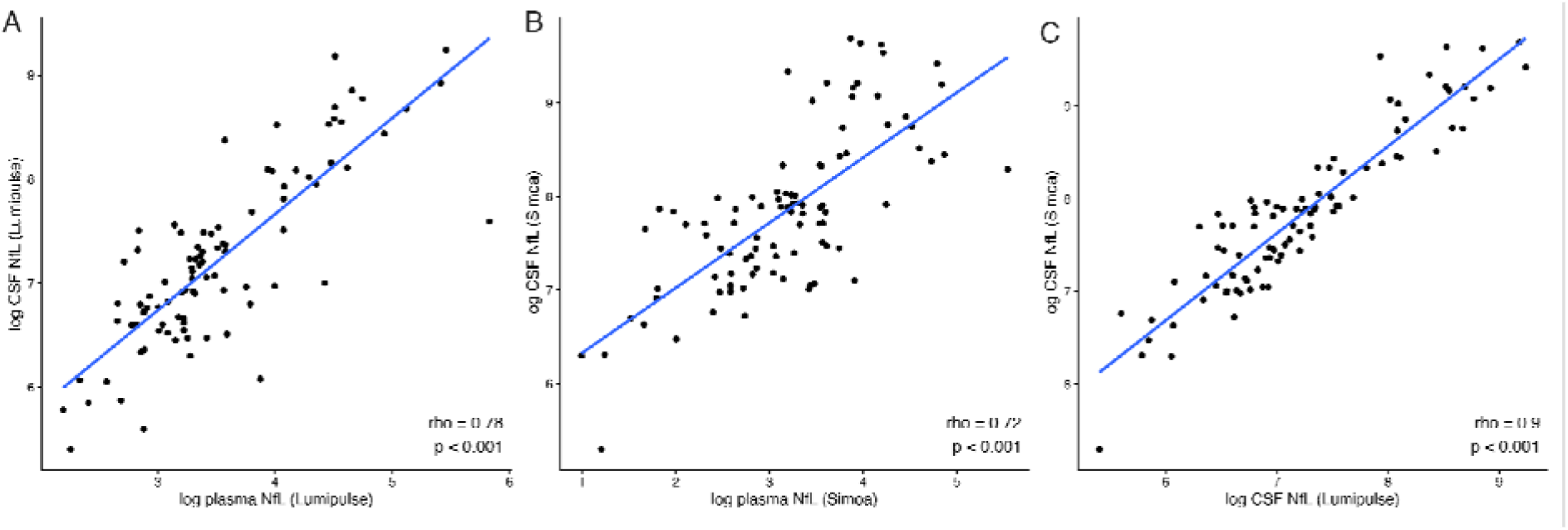
Association between plasma and cerebrospinal fluid (CSF) neurofilament light chain (NfL) concentrations and cross-platform concordance in CSF. Scatterplots showing the association between plasma and CSF NfL concentrations as well as the concordance between CSF NfL measurements obtained with the Lumipulse and Simoa platforms in the subset of patients with paired samples collected within a −61 to +61 day interval (n = 92). (A) Association between log-transformed Lumipulse plasma NfL and Lumipulse CSF NfL concentrations, (B) Association between log-transformed Simoa plasma NfL and Simoa CSF NfL concentrations, (C) Concordance between log-transformed CSF NfL concentrations measured by Lumipulse and Simoa. Spearman’s rank correlation coefficient (ρ) and corresponding p values are reported in each panel. The solid line represents the least-squares regression fit for visualization purposes.

### Plasma NfL across diagnostic categories

Plasma NfL distributions across diagnostic categories showed a broadly similar pattern on the two analytical platforms, with lower values in healthy controls and higher values in neurological groups (Figure 6; raw values in Supplementary Figure 6).

**Figure 6.**
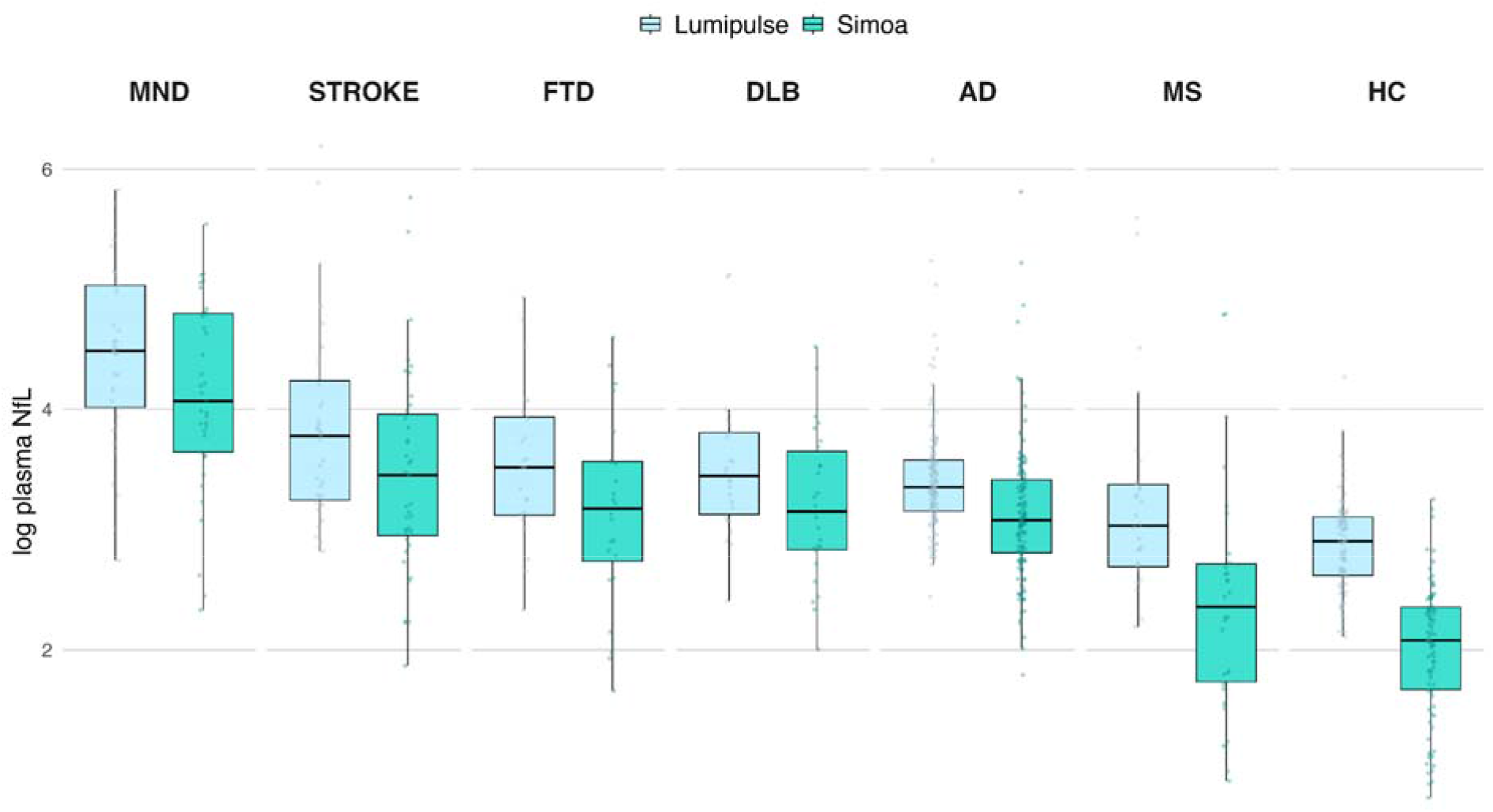
Distribution of log-transformed plasma neurofilament light chain (NfL) concentrations across diagnostic categories measured by the Lumipulse and Simoa platforms. Boxplots indicate the median and interquartile range, with individual data points overlaid. MND, motor neuron disease; STROKE, ischemic stroke; DLB, dementia with Lewy bodies; FTD, frontotemporal dementia; AD, Alzheimer’s disease; MS, multiple sclerosis; HC, healthy controls.

In permutation-based ANCOVA models including the full cohort, both diagnosis and age were significantly associated with log-transformed plasma NfL concentrations for Lumipulse (diagnosis: F(6,369) = 3.95, p<0.001; age: F(1,369) = 7.54, p = 0.007) and Simoa (diagnosis: F(6,369) = 8.21, p<0.001; age: F(1,369) = 9.70, p = 0.002). The diagnosis-by-age interaction was not significant for Lumipulse (F(6,369) = 1.64, p = 0.1376), whereas it was significant for Simoa (F(6,369) = 3.61, p = 0.0030). In a sensitivity analysis excluding healthy controls, the diagnosis-by-age interaction was no longer significant for either platform (Lumipulse: F(5,283) = 1.45, p = 0.2102; Simoa(5,283): F = 1.81, p = 0.1122). In this restricted analysis, diagnosis was not significantly associated with plasma NfL concentrations (Lumipulse(5,283): F = 1.94, p = 0.0930; Simoa: F(5,283) = 1.65, p = 0.1486), whereas age remained significant for both Lumipulse (F = 4.50, p = 0.0372) and Simoa (F = 4.63, p = 0.0304).

In the multiple linear regression sensitivity model, for Simoa, age was associated with higher log-transformed plasma NfL levels (β=0.0178, *p*<0.001). Compared with healthy controls, all diagnostic groups showed significantly higher plasma NfL concentrations, including AD (β=0.897, *p*<0.001), DLB (β=0.884, *p*<0.001), FTD (β=0.952, *p*<0.001), MND (β=1.927, *p*<0.001), MS (β=0.889, *p*<0.001), and stroke (β=1.202, *p*<0.001). Similarly, for Lumipulse, age remained associated with higher log-transformed plasma NfL levels (β=0.0113, *p*<0.001), and all diagnostic groups had significantly higher plasma NfL concentrations than healthy controls: AD (β=0.411, *p*<0.001), DLB (β=0.460, *p*<0.001), FTD (β=0.543, *p*<0.001), MND (β=1.473, *p*<0.001), MS (β=0.644, *p*<0.001), and stroke (β=0.803, *p*<0.001).

## Discussion

In this study, we investigated the analytical and clinical performances of plasma neurofilament light chain (NfL) measured with two immunoassay platforms, Simoa and Lumipulse, in a large and clinically heterogeneous cohort including healthy controls and patients with neurodegenerative, neuroinflammatory, and cerebrovascular disorders.

Plasma NfL concentrations measured with Lumipulse and Simoa were strongly correlated, in line with previous cross-platform studies, supporting the robustness of plasma NfL as a cross-platform biomarker of non-specific axonal injury [14,32–34]. However, Passing–Bablok analysis showed a systematic inter-assay difference, with both proportional and constant positive bias of Lumipulse relative to Simoa, consistent with recent head-to-head comparisons [32], while Bland–Altman analysis confirmed a significant mean bias. Thus, although the two platforms provide highly concordant individual rankings, absolute concentrations are not directly interchangeable. This has clear clinical implications, as shared cut-offs across platforms may lead to systematic misclassification and support the need for assay-specific reference ranges or conversion equations.

The Lumipulse CSF and plasma assays demonstrated excellent analytical robustness and high stability based on the 5×5 precision protocol. The Lumipulse low intra-day and inter-day CV aligns with international standards and the performances of other markers tested with the same platform [20,35] . The absence of systematic bias between replicates and the low RMS-estimated imprecision ensure that NfL measurements in CSF remain robust across a wide dynamic range and confirm the platform’s reliability for accurate clinical classification and longitudinal patient monitoring with a cumulative Total Error of 16.64%

Another relevant finding is the significant correlation between plasma and CSF NfL concentrations observed by both analytical platforms in the subset of patients with different neurodegenerative and inflammatory conditions with paired samples. In line with previous evidence showing moderate to strong CSF–blood associations [11,36,37], it supports the biological coupling between central neuroaxonal injury and peripheral NfL release, reinforcing the pathophysiological relevance of plasma NfL as a minimally invasive surrogate of CNS damage. Nevertheless, the correlation was not perfect, likely reflecting the influence of factors affecting plasma NfL dynamics, that need to be further address in larger on-going validation studies [38–41].

In addition, CSF NfL concentrations measured using Lumipulse and Simoa were very strongly correlated, suggesting that both assays capture similar inter-individual variation in the central compartment. This finding further supports the robustness of NfL quantification across different immunoassay technologies.

In the age-adjusted analyses, both diagnostic group and age were significantly associated with plasma NfL levels in the full cohort on both analytical platforms, supporting the clinical relevance of plasma NfL variation across disease categories.

Notably, a significant diagnosis-by-age interaction was observed, only for Simoa. However, this interaction disappeared after excluding healthy controls, and diagnosis was no longer significantly associated with plasma NfL within the neurological groups alone, whereas age remained significant on both platforms. Together, these findings suggest that the age-adjusted signal captured by both assays is the distinction between healthy controls and neurological disease, rather than a stable separation among neurological diagnostic categories. These findings were further supported by the multiple linear regression models. In both analytical platforms, age remained associated with higher plasma NfL concentrations, and all neurological diagnostic groups showed significantly higher plasma NfL levels than healthy controls after adjustment for age. These findings are consistent with the known increase in axonal injury across neurological conditions [1,2,42] and with NfL increase with age [12],and provides an internal biological validation of both assays across a wide dynamic range of concentrations.

This study has several strengths. First, the relatively large sample size and the inclusion of multiple neurological conditions allowed us to evaluate plasma NfL across a broad clinical spectrum and over a wide range of concentrations. Second, the use of complementary statistical approaches, including permutation-based age-adjusted analyses, multiple linear regression, correlation analyses, Passing–Bablok regression, and Bland–Altman analysis, provided a comprehensive assessment of both the clinical distribution of plasma NfL and the analytical relationship between platforms. Third, the availability of paired plasma and CSF samples in a subset of participants allowed biological anchoring of plasma NfL to the central compartment.

Several limitations should be acknowledged. First, although healthy controls were included as the reference group in the regression models, the healthy-control sample was smaller than the overall neurological cohort, and diagnostic groups were unbalanced in size and age distribution, which may have influenced between-group comparisons despite age-adjusted modeling. Second, the retrospective single-center design may limit generalizability across laboratories and clinical settings. Third, the subset with paired plasma and CSF measurements was relatively small and did not include the full clinical spectrum of the main cohort. Finally, plasma–CSF analyses were not adjusted for systemic confounders known to influence blood NfL concentrations, including renal function and BMI-related factors.

In conclusion, plasma NfL measured with both Lumipulse and Simoa showed a strong association and was significantly associated with both age and diagnostic group, supporting its clinical relevance across a broad spectrum of neurological conditions. Our findings indicate that Lumipulse and Simoa are not directly interchangeable for absolute plasma NfL concentrations, but they preserve highly concordant age-related and disease-versus-control biological information, supporting assay-specific and age-aware interpretation across platforms.

## Data Availability

All data produced in the present study are available upon reasonable request to the authors

## FIGURES

**Supplementary Table 1.** Repeatability test Plasma Level 1, PL1 (5 replicates X 5 testing days)

| NfL - Plasma LEVEL 1 | PL1-1 | PL1-2 | PL1-3 | PL1-4 | PL1-5 | Intraday MEAN | Intraday SD | CV(%) |
| --- | --- | --- | --- | --- | --- | --- | --- | --- |
| day 1 (pg/mL) | 21.30 | 19.87 | 20.71 | 20.05 | 21.70 | 20.73 | 0.79 | 3.79 |
| day 2 (pg/mL) | 19.53 | 19.91 | 21.54 | 21.34 | 20.83 | 20.63 | 0.88 | 4.27 |
| day 3 (pg/mL) | 20.55 | 20.89 | 19.99 | 19.51 | 19.55 | 20.09 | 0.61 | 3.04 |
| day 4 (pg/mL) | 21.40 | 21.82 | 20.31 | 20.01 | 19.03 | 20.51 | 1.12 | 5.44 |
| day 5 (pg/mL) | 19.41 | 19.71 | 19.93 | 20.19 | 19.61 | 19.77 | 0.30 | 1.52 |

Repeatability test of the Lumipulse G600II using 5 commercial QC available products (Plasma Level 1, PL1) tested in 5 different days. NfL, Neurofilament light-chain; SD, Standard Deviation; CV, Coefficient of Variation.

**Supplementary Table 2.**
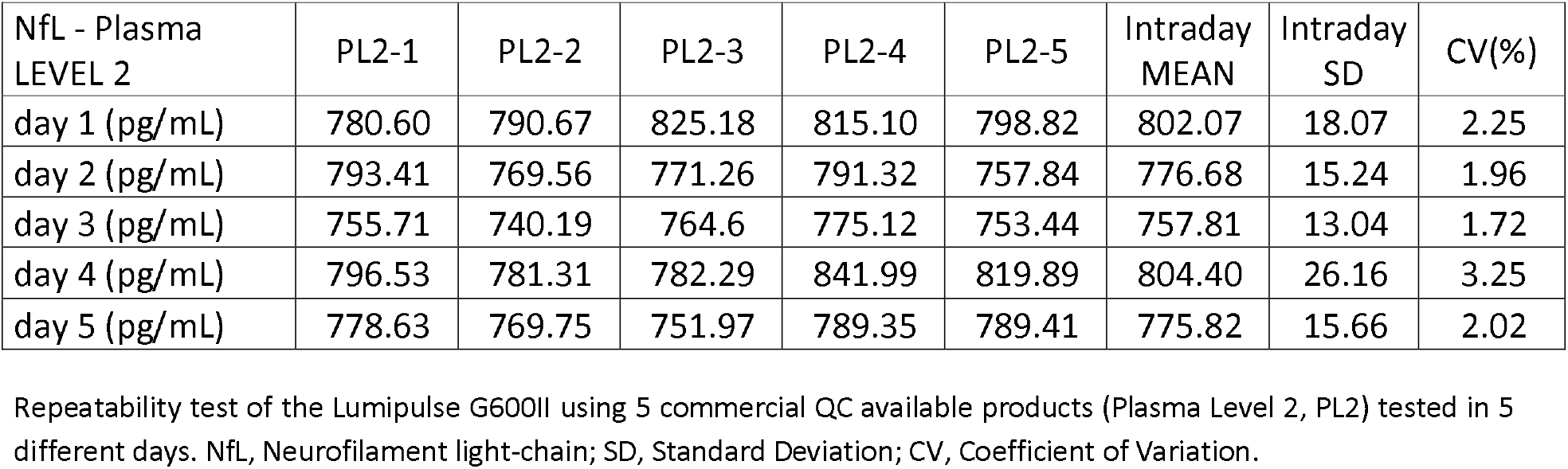
Repeatability test Plasma Level 2, PL2 (5 replicates X 5 testing days)

| NfL - Plasma LEVEL 2 | PL2-1 | PL2-2 | PL2-3 | PL2-4 | PL2-5 | Intraday MEAN | Intraday SD | CV(%) |
| --- | --- | --- | --- | --- | --- | --- | --- | --- |
| day 1 (pg/mL) | 780.60 | 790.67 | 825.18 | 815.10 | 798.82 | 802.07 | 18.07 | 2.25 |
| day 2 (pg/mL) | 793.41 | 769.56 | 771.26 | 791.32 | 757.84 | 776.68 | 15.24 | 1.96 |
| day 3 (pg/mL) | 755.71 | 740.19 | 764.6 | 775.12 | 753.44 | 757.81 | 13.04 | 1.72 |
| day 4 (pg/mL) | 796.53 | 781.31 | 782.29 | 841.99 | 819.89 | 804.40 | 26.16 | 3.25 |
| day 5 (pg/mL) | 778.63 | 769.75 | 751.97 | 789.35 | 789.41 | 775.82 | 15.66 | 2.02 |

Repeatability test of the Lumipulse G600II using 5 commercial QC available products (Plasma Level 2, PL2) tested in 5 different days. NfL, Neurofilament light-chain; SD, Standard Deviation; CV, Coefficient of Variation.

**Supplementary Table 3.** Precision results of the CSF (CSF level 1, CL1; CSF level 2, CL2; CSF level 3.

CL3) and Plasma (plasma level 1, PL1; plasma level 2, PL2) commercial QC
| Precision | CL1 | CL2 | CL3 | PL1 | PL2 |
| --- | --- | --- | --- | --- | --- |
| General mean (pg/mL) | 773.80 | 4184.24 | 19871.04 | 20.35 | 783.36 |
| DS Laboratory | 25.89 | 112.186 | 518.46 | 0.81 | 25.54 |
| CV within Laboratory (%) | 3.35 | 2.68 | 2.61 | 3.99 | 3.26 |
| CV between run (%) | 3.41 | 1.40 | 2.02 | 3.87 | 2.32 |

Summary of mean concentrations (pg/mL), laboratory standard deviations (SD), within-laboratory Coefficient of Variation (CV%), and between-run CV% evaluated for three CSF control levels (CSF Level 1, CL1; CSF Level 2, CL2; CSF Level 3, CL3) and two plasma control levels (Plasma Level 1, PL1; Plasma Level 2, PL2).

**Supplementary Table 4.** Repeatability test CSF Level 1, CL1 (5 replicates X 5 testing days)

| NfL - CSF LEVEL 1 | CL1-1 | CL1-2 | CL1-3 | CL1-4 | CL1-5 | Intraday MEAN | Intraday SD | CV(%) |
| --- | --- | --- | --- | --- | --- | --- | --- | --- |
| day 1 (pg/mL) | 779.00 | 800.00 | 760.00 | 771.00 | 772.00 | 776.40 | 14.84 | 1.91 |
| day 2 (pg/mL) | 799.00 | 810.00 | 794.00 | 760.00 | 747.00 | 782.00 | 27.05 | 3.46 |
| day 3 (pg/mL) | 794.00 | 759.00 | 824.00 | 766.00 | 760.00 | 780.60 | 28.14 | 3.61 |
| day 4 (pg/mL) | 744.00 | 765.00 | 753.00 | 772.00 | 838.00 | 774.40 | 37.15 | 4.79 |
| day 5 (pg/mL) | 757.00 | 778.00 | 736.00 | 770.00 | 737.00 | 755.60 | 18.98 | 2.51 |

Repeatability test of the Lumipulse G600II using 5 commercial QC available products (CSF Level 1, CL1) tested in 5 different days. NfL, Neurofilament light-chain; CSF, cerebrospinal fluid; SD, Standard Deviation; CV, Coefficient of Variation.

**Supplementary Table 5.** Repeatability test CSF Level 2, CL2 (5 replicates X 5 testing days)

| NfL - CSF LEVEL 2 | CL2-1 | CL2-2 | CL2-3 | CL2-4 | CL2-5 | Intraday MEAN | Intraday SD | CV(%) |
| --- | --- | --- | --- | --- | --- | --- | --- | --- |
| day 1 (pg/mL) | 4266.00 | 4317.00 | 4157.00 | 4227.00 | 4231.00 | 4239.60 | 58.62 | 1.38 |
| day 2 (pg/mL) | 4269.00 | 4375.00 | 4352.00 | 4208.00 | 4166.00 | 4274.00 | 89.90 | 2.10 |
| day 3 (pg/mL) | 4022.00 | 3960.00 | 4098.00 | 4072.00 | 4062.00 | 4042.80 | 53.75 | 1.33 |
| day 4 (pg/mL) | 4204.00 | 4265.00 | 4253.00 | 4312.00 | 4187.00 | 4244.20 | 49.99 | 1.18 |
| day 5 (pg/mL) | 4112.00 | 4149.00 | 4104.00 | 4077.00 | 4161.00 | 4120.60 | 34.24 | 0.83 |

Repeatability test of the Lumipulse G600II using 5 commercial QC available products (CSF Level 2, L2) tested in 5 different days. NfL, Neurofilament light-chain; CSF, cerebrospinal fluid; SD, Standard Deviation; CV, Coefficient of Variation.

**Supplementary Table 6.**
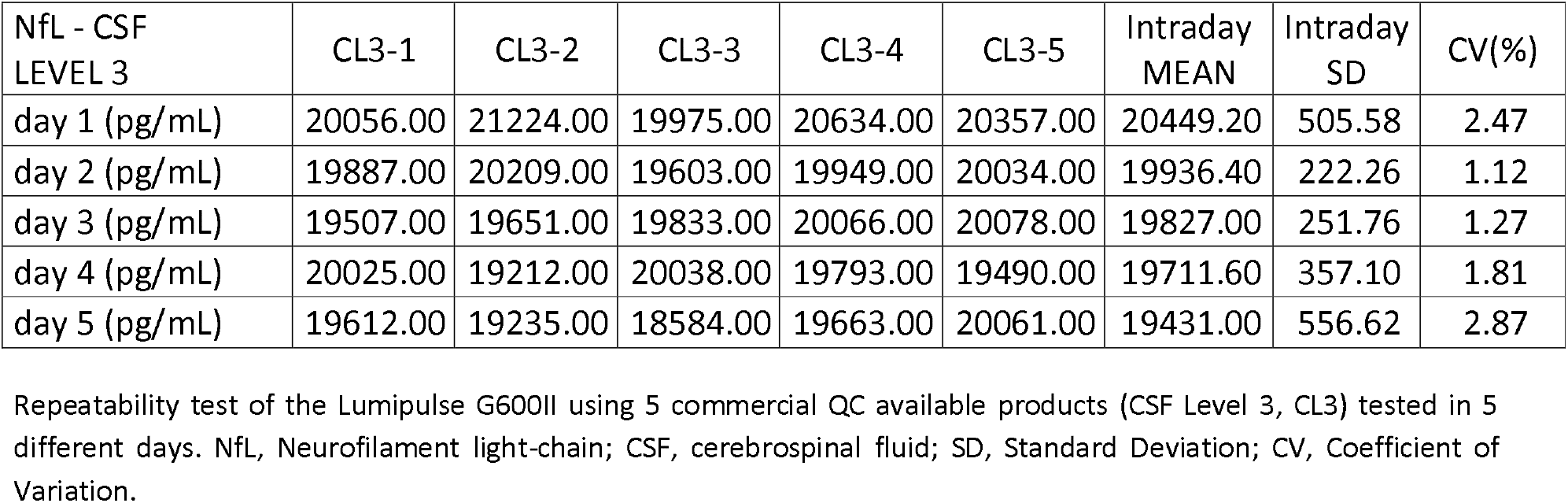
Repeatability test CSF Level 3, CL3 (5 replicates X 5 testing days)

| NfL - CSF LEVEL 3 | CL3-1 | CL3-2 | CL3-3 | CL3-4 | CL3-5 | Intraday MEAN | Intraday SD | CV(%) |
| --- | --- | --- | --- | --- | --- | --- | --- | --- |
| day 1 (pg/mL) | 20056.00 | 21224.00 | 19975.00 | 20634.00 | 20357.00 | 20449.20 | 505.58 | 2.47 |
| day 2 (pg/mL) | 19887.00 | 20209.00 | 19603.00 | 19949.00 | 20034.00 | 19936.40 | 222.26 | 1.12 |
| day 3 (pg/mL) | 19507.00 | 19651.00 | 19833.00 | 20066.00 | 20078.00 | 19827.00 | 251.76 | 1.27 |
| day 4 (pg/mL) | 20025.00 | 19212.00 | 20038.00 | 19793.00 | 19490.00 | 19711.60 | 357.10 | 1.81 |
| day 5 (pg/mL) | 19612.00 | 19235.00 | 18584.00 | 19663.00 | 20061.00 | 19431.00 | 556.62 | 2.87 |

Repeatability test of the Lumipulse G600II using 5 commercial QC available products (CSF Level 3, CL3) tested in 5 different days. NfL, Neurofilament light-chain; CSF, cerebrospinal fluid; SD, Standard Deviation; CV, Coefficient of Variation.

**Supplementary Table 7.**
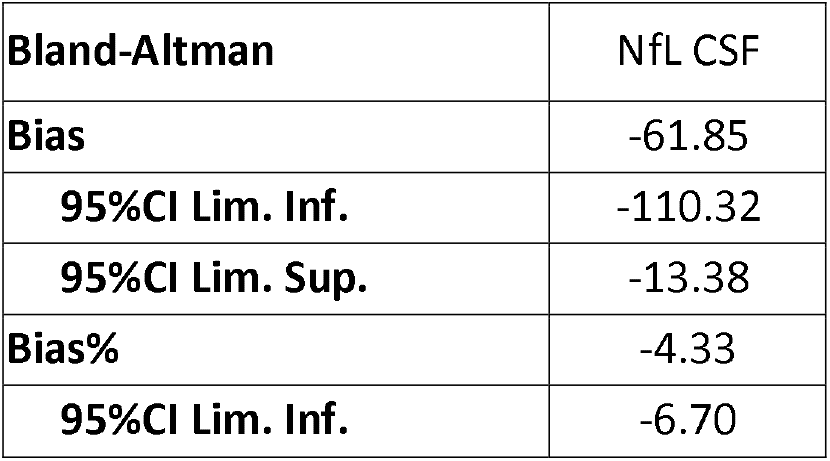
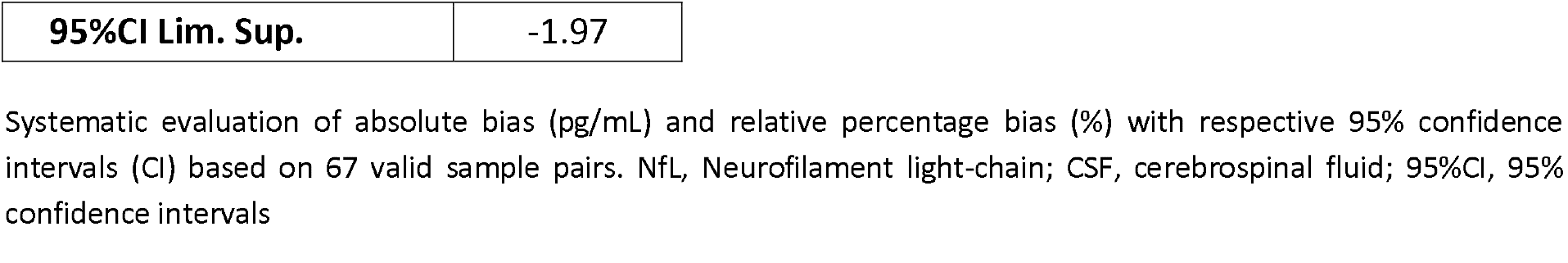
Bland-Altman analysis for NfL analyzed in double repetitions.

| Bland-Altman | NfL CSF |
| --- | --- |
| Bias | -61.85 |
| 95%CI Lim. Inf. | -110.32 |
| 95%CI Lim. Sup. | -13.38 |
| Bias% | -4.33 |
| 95%CI Lim. Inf. | -6.70 |
| **95%CI Lim. Sup.** | -1.97 |

Systematic evaluation of absolute bias (pg/mL) and relative percentage bias (%) with respective 95% confidence intervals (CI) based on 67 valid sample pairs. NfL, Neurofilament light-chain; CSF, cerebrospinal fluid; 95%CI, 95% confidence intervals

**Supplementary Table 8.** Passing-Bablok regression for NfL analyzed in double repetitions.

|  |  |
| --- | --- |
| **Passing-Bablok regression** | NfL CSF |
| **Slope** | 1.02 |
| **95%CI Lim. Inf.** | 0.99 |
| **95%CI Lim. Sup.** | 1.06 |
| **Intercept** | 19.58 |
| **95%CI Lim. Inf.** | -10.57 |
| **95%CI Lim. Sup.** | 50.00 |

Method comparison statistics showing slope and intercept estimates with corresponding 95% confidence intervals (CI) to evaluate proportional and constant systematic deviations across 67 sample pairs. NfL, Neurofilament light-chain; CSF, cerebrospinal fluid; 95%CI, 95% confidence intervals

**Supplementary Figure 1.**
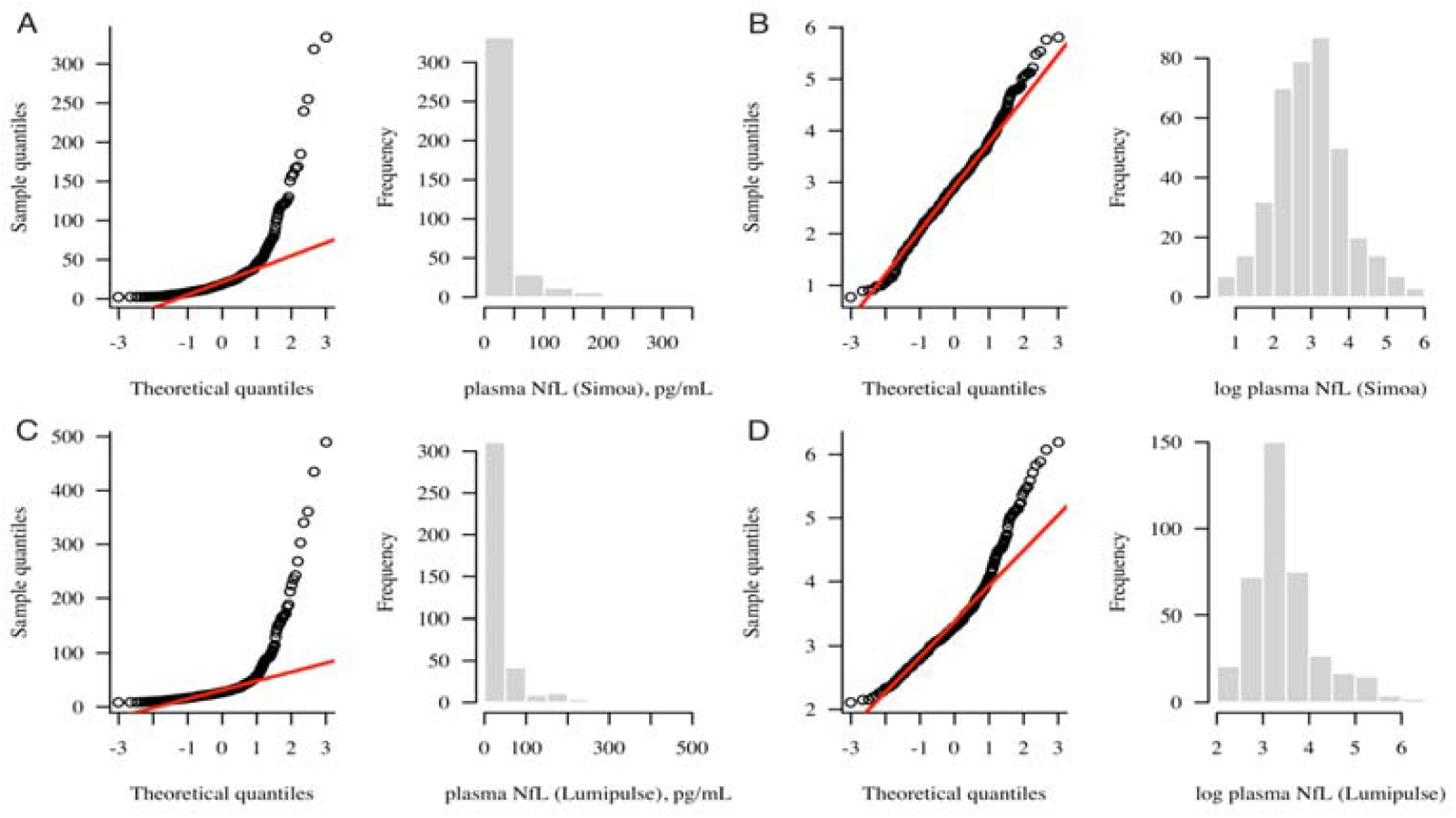
Distribution of plasma neurofilament light chain (NfL) concentrations measured by Simoa and Lumipulse. (A) Q–Q plot and histogram of raw plasma NfL values measured by Simoa. (B) Q–Q plot and histogram of log-transformed plasma NfL values measured by Simoa. (C) Q–Q plot and histogram of raw plasma NfL values measured by Lumipulse. (D) Q–Q plot and histogram of log-transformed plasma NfL values measured by Lumipulse.

**Supplementary Figure 2.**
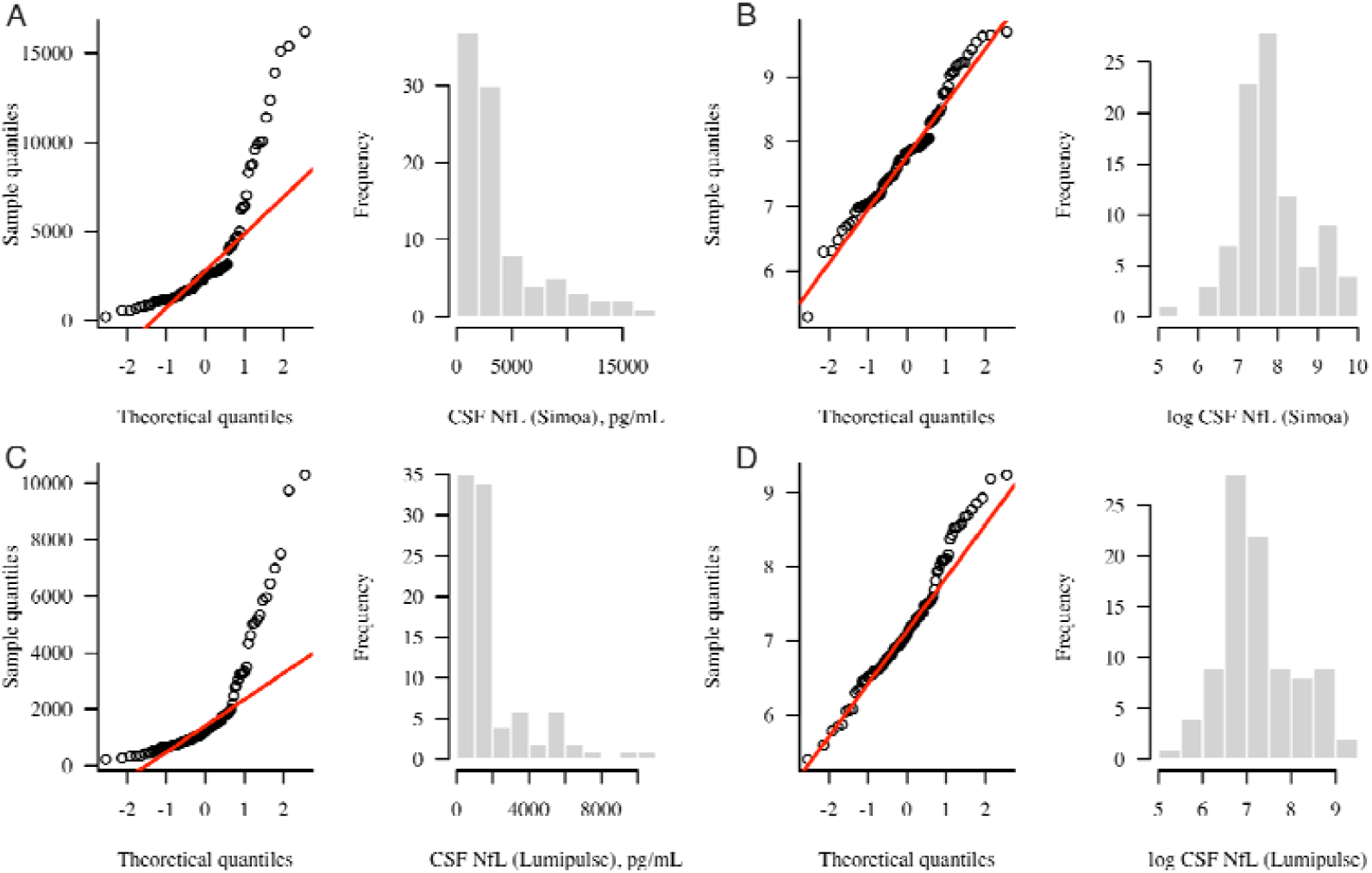
Distribution of CSF neurofilament light chain (NfL) concentrations measured by Simoa and Lumipulse. (A) Q–Q plot and histogram of raw CSF NfL values measured by Simoa. (B) Q–Q plot and histogram of log-transformed CSF NfL values measured by Simoa. (C) Q–Q plot and histogram of raw CSF NfL values measured by Lumipulse. (D) Q– Q plot and histogram of log-transformed CSF NfL values measured by Lumipulse.

**Supplementary Figure 3:**
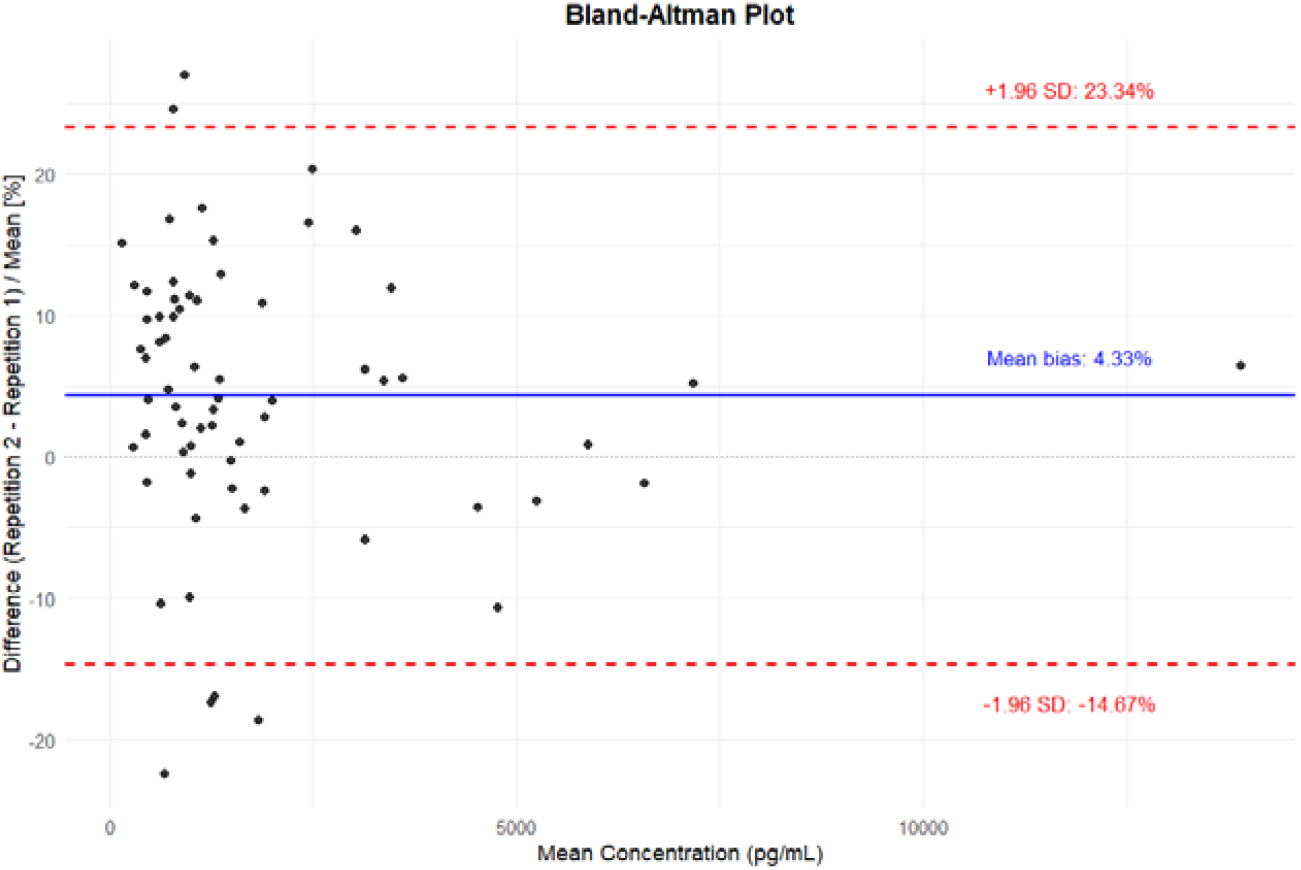
Bland-Altman plot of 67 CSF specimens analyzed in double repetitions. The relative percentage differences between repetitions are plotted against the mean concentration of the replicates (pg/mL). The solid blue line indicates the mean systematic bias (-4.33%, 95% CI: -6.70% to -1.97%), and the horizontal dotted line marks the zero-bias reference. The red dashed lines define the 95% Limits of Agreement (23.34% to -14.67%, representing ±1.96 SD).

**Supplementary Figure 4:**
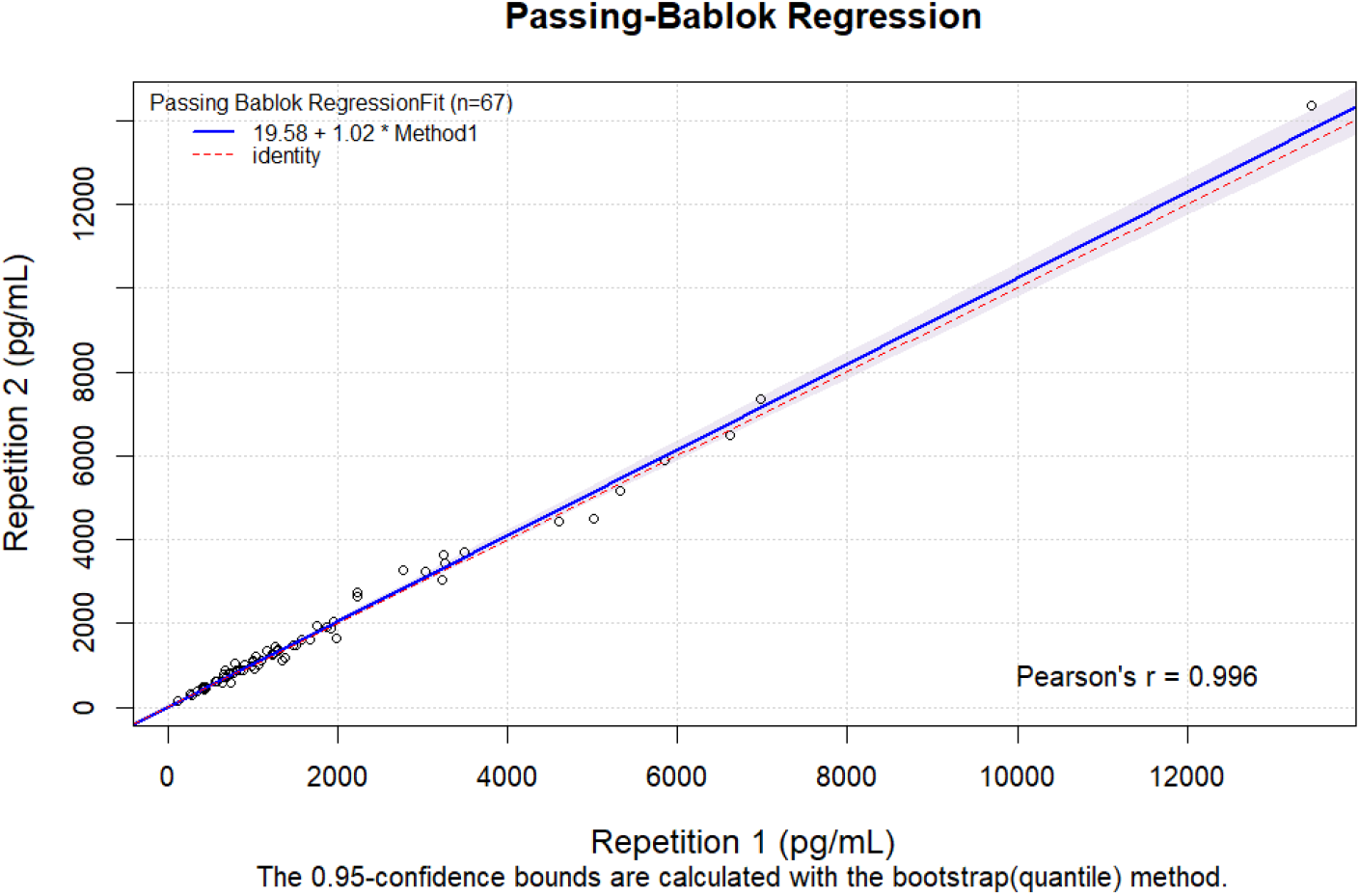
Passing-Bablok of 67 CSF specimens analyzed in double repetitions. Passing–Bablok regression of CSF NfL concentrations within 67 CSF specimens tested in double repetitions. The solid line represents the regression fit, and the dashed red line indicates the line of identity (y = x).

**Supplementary Figure 5.**
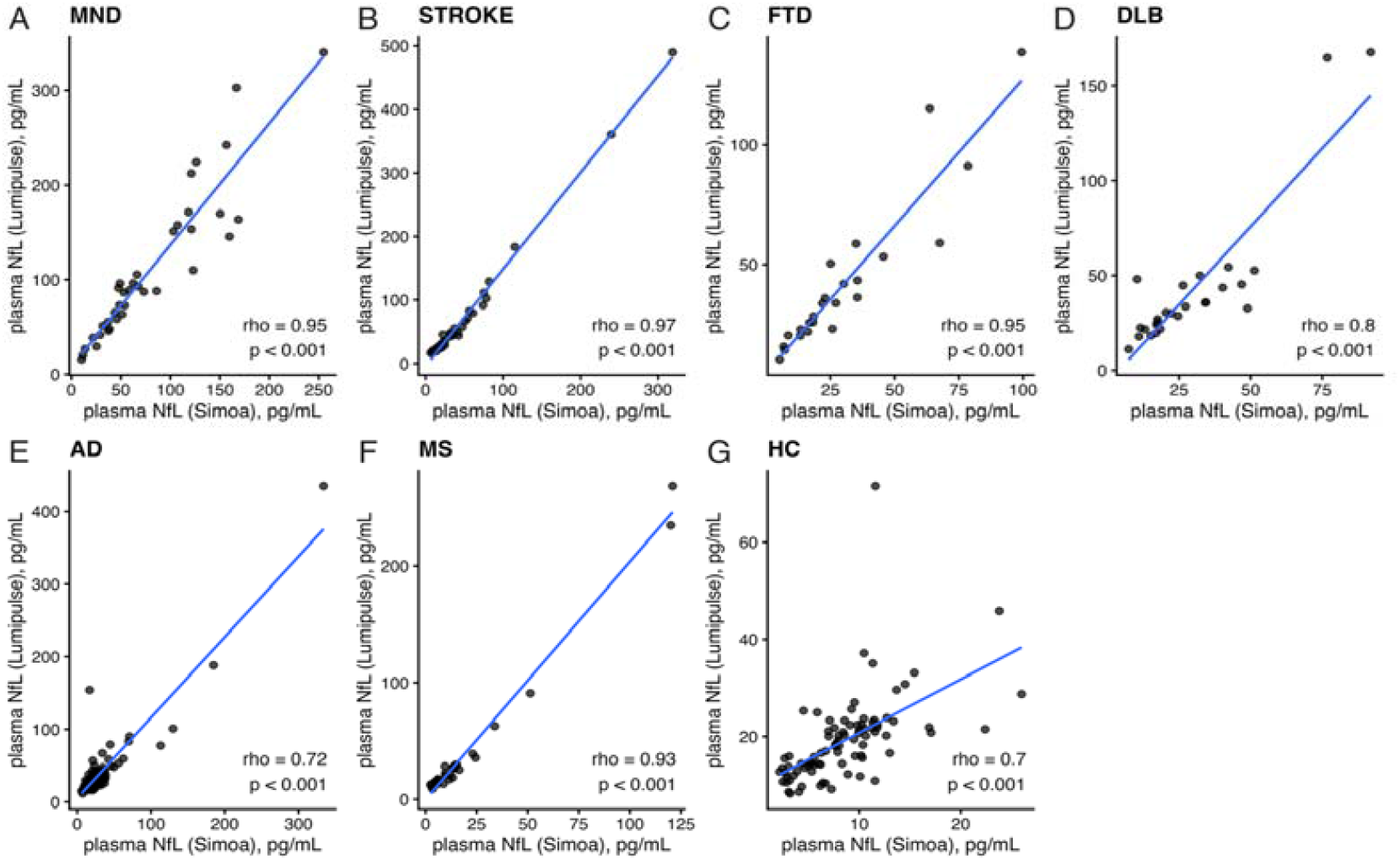
Spearman correlation between plasma neurofilament light chain (NfL) measured by Lumipulse and Simoa within diagnostic categories (raw values) Multipanel scatterplots showing the association between raw plasma NfL concentrations (pg/mL) measured by Lumipulse and Simoa in (A) motor neuron disease (MND), (B) ischemic stroke (STROKE), (C) frontotemporal dementia (FTD), (D) dementia with Lewy bodies (DLB), (E) Alzheimer’s disease (AD), (F) multiple sclerosis (MS), and (G) healthy controls (HC). Spearman’s rank correlation coefficient (ρ) and p value are reported within each panel; the solid line indicates the least-squares fit for visualization.

**Supplementary Figure 6.**
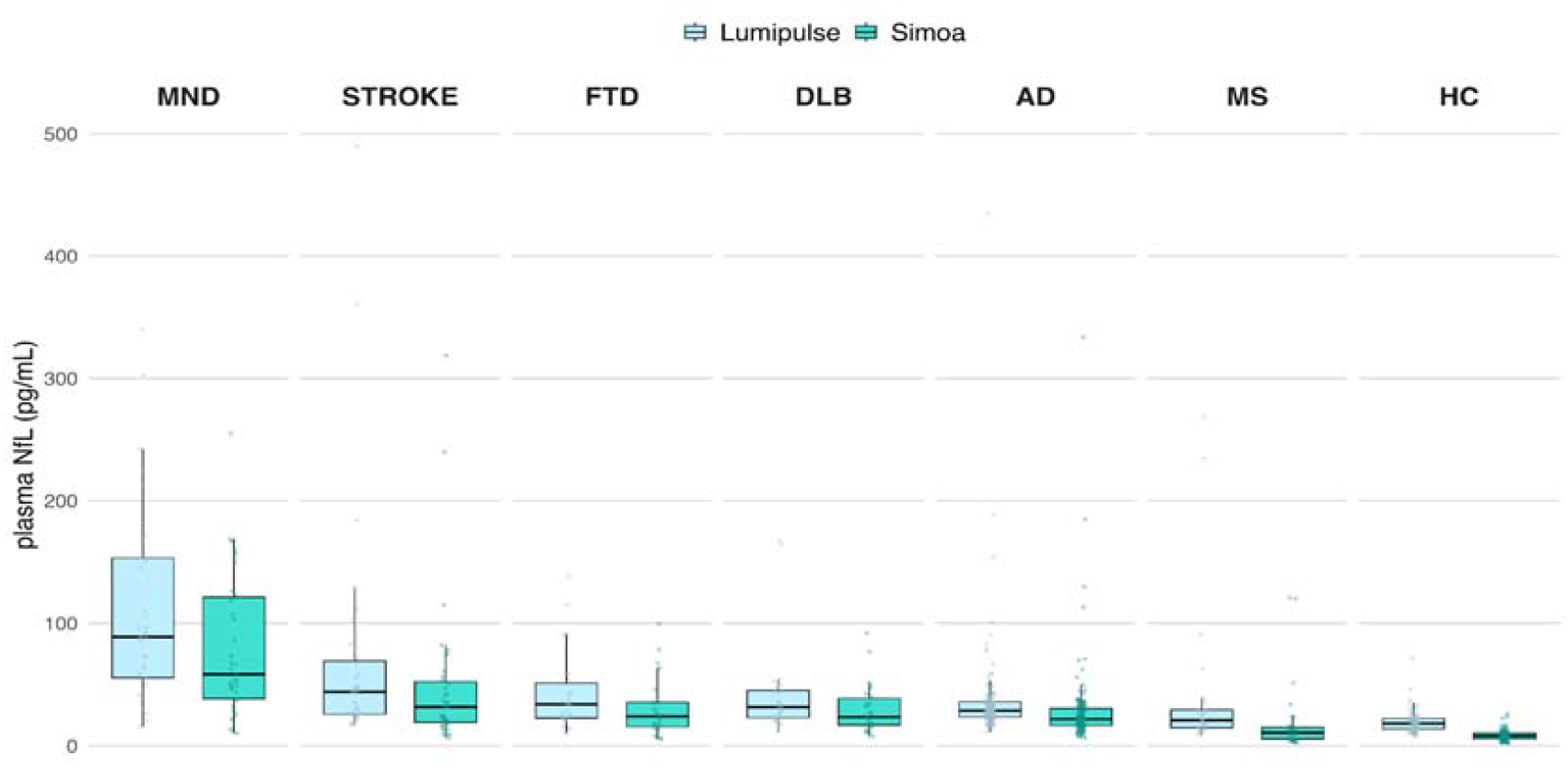
Distribution of plasma neurofilament light chain (NfL) raw concentrations across diagnostic categories measured by the Lumipulse and Simoa platforms. Boxplots indicate the median and interquartile range, with individual data points overlaid. MND, motor neuron disease; STROKE, ischemic stroke; FTD, frontotemporal dementia; DLB, dementia with Lewy bodies; AD, Alzheimer’s disease; MS, multiple sclerosis; HC, healthy controls.

## Acknowledgment

The study has been supported by the European Union -Next Generation EU - NRRP M6C2 - Investment 2.1 Enhancement and strengthening of biomedical research in the NHS– PNRR-Health PNRR-MAD-2022-12376110.

The authors wish to thank the study participants who took part in this research. We are very grateful for the assistance received from the Neurology department of the Spedali Civili of Brescia.

The authors would like also to express their sincere gratitude to Dr. Sara Baldelli and Dr. Marco Chiarini for their insightful contributions to our understanding of the analytical aspects of this work, and their great dedication to translational research in laboratory medicine and patient care, in both chronic and acute clinical settings, including around-the-clock emergency support.

## Funding declarations

Andrea Toja declares no funding.

Virginia Quaresima declares no funding.

Chiara Tolassi has been supported by the PNRR-Health PNRR-MAD-2022-12376110

Tommaso Merati declares no funding.

Chiara Trasciatti has been supported by the H2020 IMI IDEA-FAST (ID853981) project

Simona G. Signorini, Andrea Morotti, Francesco Berinato, Loris Poli, Liberato Stabile, Irene GIrotto, Marianna Bertoni, Cinzia Zatti, Alessandro Magliozzi, Caterina Martinuzzo, Claudia Pangrazio, Klaudia Eshja, Giulia Foresti, Ilenia Libri, Eqrem Rusi, Marta Bianchi, Viviana Cristillo, Irene Volonghi, Alice Galli, Andrea Rizzardi, Salvatore Caratozzolo, Chiara Agosti declare no funding.

Rosanna Colao has been supported by the PNRR-Health PNRR-MAD-2022-12376110

Carmelo Rodolico has been supported by the PNRR-Health PNRR-MAD-2022-12376110

Elena Marcello received funding from Italian Ministry of Research and University (MUR) (PRIN2022 PNRR P2022R2E8N to EM), from the Giovanni Armenise Harvard Foundation and AIRALZH ONLUS (2023 Armenise Harvard-AIRALZH Mid-Career Award in Neurodegenerative Diseases - AHA MCA).

Fabrizio Gardoni received funding from Italian Ministry of Research and University (MUR) (PRIN2022YY85P5 to EM).

MDL received funding from Italian Ministry of Research and University (MUR) (PRIN2022 PNRR P2022TKN8C, Fondo Italiano per la Scienza FIS00000560 - Stone to MDL).

HZ is a Wallenberg Scholar and a Distinguished Professor at the Swedish Research Council supported by grants from the Swedish Research Council (#2023-00356, #2022-01018 and #2019-02397), the European Union’s Horizon Europe research and innovation programme under grant agreement No 101053962, Swedish State Support for Clinical Research (#ALFGBG-71320), the Alzheimer Drug Discovery Foundation (ADDF), USA (#201809-2016862), the AD Strategic Fund and the Alzheimer’s Association (#ADSF-21-831376-C, #ADSF-21-831381-C, #ADSF-21-831377-C, and #ADSF-24-1284328-C), the European Partnership on Metrology, co-financed from the European Union’s Horizon Europe Research and Innovation Programme and by the Participating States (NEuroBioStand, #22HLT07), the Bluefield Project, Cure Alzheimer’s Fund, the Olav Thon Foundation, the Erling-Persson Family Foundation, Familjen Rönströms Stiftelse, Familjen Beiglers Stiftelse, Stiftelsen för Gamla Tjänarinnor, Hjärnfonden, Sweden (#FO2022-0270), the European Union’s Horizon 2020 research and innovation programme under the Marie Skłodowska-Curie grant agreement No 860197 (MIRIADE), the European Union Joint Programme – Neurodegenerative Disease Research (JPND2021-00694), the National Institute for Health and Care Research University College London Hospitals Biomedical Research Centre, the UK Dementia Research Institute at UCL (UKDRI-1003), and an anonymous donor.

Nicholas J Ashton declares no funding.

Duilio Brugnoni declares no funding.

APi has been supported by grants of Airalzh Foundation AGYR2021 Life-Bio Grant, The LIMPE-DISMOV Foundation Segala Grant 2021, the Italian Ministry of University and Research PRIN COCOON (2017MYJ5TH), PRIN 2022PNJS5Z and PRIN PNRR (P20224ZHM9), DIGI-BRAIN the H2020 IMI IDEA-FAST (ID853981), Italian Ministry of Health, Grant/Award Number: RF-2018-12366209, PNRR-Health PNRR-MAD-2022-12376110 and PNRR-MCNT2-2023-12378387, The MJFF Foundation Grant 022343, #NEXTGENERATIONEU (NGEU) funded by the Ministry of University and Research (MUR), National Recovery and Resilience Plan (NRRP), project MNESYS (PE0000006) – a multiscale integrated approach to the study of the nervous system in health and disease (DN. 1553 11.10.2022) – subproject DIGI-BRAIN; the European Union –

APa has been supported by grants of the Italian Ministry of University and Research PRIN COCOON (2017MYJ5TH) and PRIN 2021 RePlast (20202THZAW), Prin 2022 EGADi (P2022TKN8C) the H2020 IMI IDEA-FAST (ID853981), #NEXTGENERATIONEU (NGEU) funded by the Ministry of University and Research (MUR), National Recovery and Resilience Plan (NRRP), project MNESYS (PE0000006) – a multiscale integrated approach to the study of the nervous system in health and disease (DN. 1553 11.10.2022) – subproject DIGI-BRAIN Grant/Award Number: RF-2018-12366209, RF-2019-12369272 and PNRR-Health PNRR-MAD-2022-12376110, the Next Generation EU - NRRP M6C2 - Investment 2.1 Enhancement and strengthening of biomedical research in the NHS-PNRR – PNRR PNRR-Health PNRR-MAD-2022-12376110; PRIN 2020 Prot. 20202THZAW “RE-Plast: targeting functional and structural plasticity in Alzheimer disease. From diagnosis to treatment” and PRIN 2022 “P2022TKN8C” Extracellular network and related Genetic underpinnings as a hub in Alzheimer’s Disease (EGADi).

## Author contributions

AT, VQ, DB, Api and APa contributed to the conception and design of the study. AT and VQ performed the data analysis and prepared the figures. VQ, CT, SGS, IG contributed to the biological analyses. TM, CT, AM, FB, LP, LS, CM, ER, MB, VC, AG AR, SC, RC, CR contributed to data acquisition. AT, VQ, CT, EM, FG, MdL, HZ, NJA, DB, Api and ApA contributed to drafting and revising the manuscript.

## Competing interests

Andrea Toja declares no conflict of interest

Virginia Quaresima declares no conflict of interest

Chiara Tolassi declares no conflict of interest

Tommaso Merati declares no conflict of interest

Chiara Trasciatti declares no conflict of interest

Simona G. Signorini declares no conflict of interest

Andrea Morotti declares no conflict of interest

Francesco Berinato declares no conflict of interest

Loris Poli declares no conflict of interest

Liberato Stabile declares no conflict of interest

Irene Girotto declares no conflict of interest

Marianna Bertoni declares no conflict of interest

Cinzia Zatti declares no conflict of interest

Alessandro Magliozzi declares no conflict of interest

Caterina Martinuzzo declares no conflict of interest

Claudia Pangrazio declares no conflict of interest

Klaudia Eshja declares no conflict of interest

Giulia Foresti declares no conflict of interest

Ilenia Libri declares no conflict of interest

Eqrem Rusi declares no conflict of interest

Marta Bianchi declares no conflict of interest

Viviana Cristillo declares no conflict of interest

Irene Volonghi declares no conflict of interest

Alice Galli declares no conflict of interest

Andrea Rizzardi declares no conflict of interest

Salvatore Caratozzolo declares no conflict of interest

Chiara Agosti declares no conflict of interest

Rosanna Colao declares no conflict of interest

Carmelo Rodolico declares no conflict of interest

Elena Marcello received speaker fees from Eli Lilly and GE Healthcare, advisory board fees from Roche, teaching fees from Eisai

Fabrizio Gardoni received advisory board and speaker fees from Eli Lilly. Monica di Luca received advisory board fees from Roche

Henrik Zetterberg has served at scientific advisory boards and/or as a consultant for Abbvie, Acumen, Alector, Alzinova, ALZpath, Amylyx, Annexon, Apellis, Artery Therapeutics, AZTherapies, Cognito Therapeutics, CogRx, Denali, Eisai, Enigma, LabCorp, Merck Sharp & Dohme, Merry Life, Nervgen, Novo Nordisk, Optoceutics, Passage Bio, Pinteon Therapeutics, Prothena, Quanterix, Red Abbey Labs, reMYND, Roche, Samumed, ScandiBio Therapeutics AB, Siemens Healthineers, Triplet Therapeutics, and Wave, has given lectures sponsored by Alzecure, BioArctic, Biogen, Cellectricon, Fujirebio, LabCorp, Lilly, Novo Nordisk, Oy Medix Biochemica AB, Roche, and WebMD, is a co-founder of Brain Biomarker Solutions in Gothenburg AB (BBS), which is a part of the GU Ventures Incubator Program, and is a shareholder of CERimmune Therapeutics (outside submitted work).

Nicholas Ashton declares no conflict of interest

Duilio Brugnoni declares no conflict of interest

Andrea Pilotto received consultancy/speaker fees from Abbvie, Angelini, Bial, Eli Lilly, Lundbeck, Roche and Zambon pharmaceuticals. He acts as consultant as part of advisory Board of Angelini Pharma and BIAL pharmaceutics.

Alessandro Padovani received personal compensation as a consultant/scientific advisory board member for Biogen, Eisai Eli Lilly, General Healthcare (GE), Lundbeck, Nestlè, Roche.

## Notes

### Author Declarations

Ethics Committee of ASST Spedali Civili di Brescia gave ethical approval for this work (NP 1471, DMA, Brescia; Neuromultibio Study NP5285, approved on 10 May 2022). The study was performed in conformity with the Declaration of Helsinki; informed consent was obtained from each study participant or their legally authorized representative.

